# Endovascular endothelial cell biopsy: a systematic review

**DOI:** 10.1101/2025.04.08.25325399

**Authors:** Charles Piercy, John Adam, James N Harrison, Christian Heiss, Paola Campagnolo, Ben Creagh-Brown

## Abstract

**Background:** Endovascular endothelial cell biopsy (ECBx) allows direct sampling of endothelial cells (ECs) and subsequent assessment of EC function in health and disease. Our systematic review aims to summarise the current literature describing protocols to obtain and analyse EC and obtain pooled estimates of success rate and EC numbers obtained with different techniques in different populations.

**Methods:** Studies were identified on Medline using the terms ‘endothelial cell’ AND ‘biopsy’ AND ‘humans’. All primary research involving EC biopsy was included, while animal studies or non-primary research studies were excluded from the review. The data was integrated into a narrative review and the study was registered with PROSPERO (CRD42022278551).

**Results:** Of all journal articles identified (n=784), 51 articles were included in this review. Most studies (n=46) used a J wire method for EC sampling from blood vessels in the arm (superficial arm veins [n=30], brachial artery [n=3], radial artery [n=4]) while others used devices including stent retrievers (n=1) and endovascular coils (n=2). The pooled success rate was 91±15%. The average yield was 1,058±893 cells per biopsy, with variability depending on procedural factors such as wire placement depth and technique. No significant complications were reported. Several analytical techniques were used to evaluate the isolated ECs with the most common technique being immunofluorescence (n=36) and only a few studies reported *ex vivo* culture of the cells. Risk of bias assessment and statistical analysis not performed due to heterogeneity of data and variability in reporting.

**Conclusion:** Endothelial cells can be obtained with a variety of techniques with a high success rate and minimal complications. This review highlights novel research opportunities provided by ECBx.

## Introduction

The vascular endothelium is a cellular monolayer lining all blood vessels and it possesses diverse physiological functions, including modulation of vascular tone, regulation of thrombosis, maintenance of vascular integrity, and influence on leukocyte adhesion and migration(1). Endothelial dysfunction is well recognised as playing a critical role in classic vascular diseases such as hypertension and atherosclerosis, as well as in a variety of other pathologies(2). Given their central role in these conditions, the study of endothelial cells (EC) is essential for advancing our understanding of vascular disorders.

There are a range of techniques available for the evaluation of endothelial function for diagnostic or investigative purposes: i) Flow mediated dilation (FMD) is a physiological indirect measurement of nitric oxide-mediated vasodilation in response to endothelial sheer stress during reactive hyperaemia of the forearm (3); ii) venous occlusion plethysmography; iii) angiography with intra-arterial infusion of endothelium-dependent vasodilators such as acetylcholine; iv) imaging techniques include optical coherence tomography (OCT); v) near-infrared spectroscopy (NIRS); vi) laser doppler; vii) sampling of the blood for biomarkers, either cellular or non-cellular. Cellular markers include circulating endothelial cells, and endothelial progenitor cells. Non-cellular markers include mediators of endothelial cell function (angiopoietin-1 and -2, nitrite, RS/XNO)(4), components of the coagulation pathway (von Willebrand Factor, ADAMTS13, and thrombomodulin), soluble cell-surface adhesion molecules (soluble E-selectin, sICAM-1, and sVCAM-1), regulators of vascular tone and permeability (VEGF and sFlt-1)(3,5–9), or endothelial microparticles(10).

Direct sampling of EC from humans is possible via an endovascular biopsy (ECBx) technique. First described by Feng in 1999, the technique has been modified and adapted by subsequent investigators (11). Although there are a range of different approaches, common to them all is the collection of EC from an intravascular or endovascular device, typically a metal guidewire with a J-shaped tip (J-wire). To date, there have been no review of these different approaches or attempts to publish a unified, systematically assessed approach.

This review aims to analyse and present the current literature describing protocols to isolate and analyse EC from humans in vivo and generate pooled estimates of success rate and EC numbers obtained with different techniques in different populations.

## Methods

### Information sources and search strategy

One electronic database (Medline) was searched using the keywords ‘endothelial cell’ and ‘biopsy’ in ‘human’. Boolean operators used were ‘and’ with each output reviewed to identify suitable papers in keeping with the inclusion and exclusion criteria. Limits were applied to focus the review on studies between January 1996 to August 2024. This database was last searched on 08 Aug 2024. The start date was chosen to capture any earlier publications than Feng *et al* in 1999, who first reported this technique. Secondary searches included a manual review of references not originally identified during the primary search by identifying relevant entries from the bibliography of selected papers and by contacting authors of publications. The protocol for this review was submitted and reviewed by Prospero (CRD42022278551). The review was conducted following the PRISMA 2020 guidelines.

### Eligibility criteria

The focus of this paper is to review the techniques used to obtain vascular EC in humans. Inclusion criteria were 1) primary research studies on EC biology, 2) use of a recognised vascular intervention to obtain EC, 3) EC obtained from human patients and volunteers. Studies were excluded if they met any of the following criteria 1) non-human sampling, 2) not endothelial cells biopsied, 3) non-primary research, 4) manuscript not available in English.

### Selection process

The Medline output was split randomly amongst three reviewers to appraise each study and remove any considered ineligible. Studies were assessed by analysing the title and abstract, and those that were ineligible (see ‘exclusion criteria’) were documented and not included. Those that were considered eligible were reviewed again to reach a consensus on inclusion. Risk of bias was not per

### Data collection process

Each study included in this report was read independently by three reviewers. Each reviewer created a database to log information that corresponded to the research objectives. Once all reviews were completed, the databases were merged creating a final report. The data collection process did not use any form of automation to aid the process.

### Data items

The primary outcome was to identify techniques used to obtain biopsy samples, reported success rates, and number of EC retrieved. For each entry, details were collected in regard to the presenting pathologies, type of wire used, number of wires used per sampling, location of the biopsy, type of buffers used, enrichment methods, yield, and analysis. The secondary outcome was to compare the success rates between different biopsy techniques and samples sites. Success rate was defined as the number of biopsy samples that yielded enough viable cells for further analysis. Patient safety data including complications were collected. Data was then visually represented using BioRender or tabulated.

### Narrative synthesis

Results were generated using a narrative synthesis. The initial narrative synthesis involved tabulating the included studies and creating a summary of each study. The results were discussed amongst the three reviewers and then split into structured themes focusing on patient characteristics, type of wire used, number of wires used, depth of wire insertion , biopsy location and success rate. Size of wire used was either taken from the source study or if not documented, from the method the study references. Number of wires used was either taken from the source study or assigned a value of one if not documented. Where a range was given for EC yield rather than an average, the lower number was used. Summary of affects was calculated by selecting individual parameters and comparing that overall EC yield using weighted means. Only studies that listed one parameter per variable (e.g. using only one size of wire for the whole study) were selected for analysis. Absolute difference between wire size and blood vessel was calculated by first converting the diameter of the guide wire from inches to mm, then taking subtracting that from the average size of the blood vessel.

### Statistical methods

All statistical analyses were performed in RStudio (Build 2024.08.0+341) using the ggplot2 and dplyr packages. Weighted means and corresponding standard deviations were calculated for each variable. Linear regression was used to model the relationship between the size of the guidewire and the effect on yield using the dplyr package. PRISMA diagram was created using DiagrammeR package. UpSet plot was created using complexupset.

## Results

### Study selection

A total of 784 papers containing the key search terms were identified from the Medline database and other sources. All papers were then reviewed for eligibility to be included in the study. Most papers excluded were describing a different type of biopsy technique, for example tissue biopsy with EC isolation (n=571). Other reasons for exclusion are detailed in Figure 1. After screening, 51 papers were deemed suitable for inclusion into the current review. Full information about each article and data extraction can be seen in supplementary table 1.

**Figure 1.**
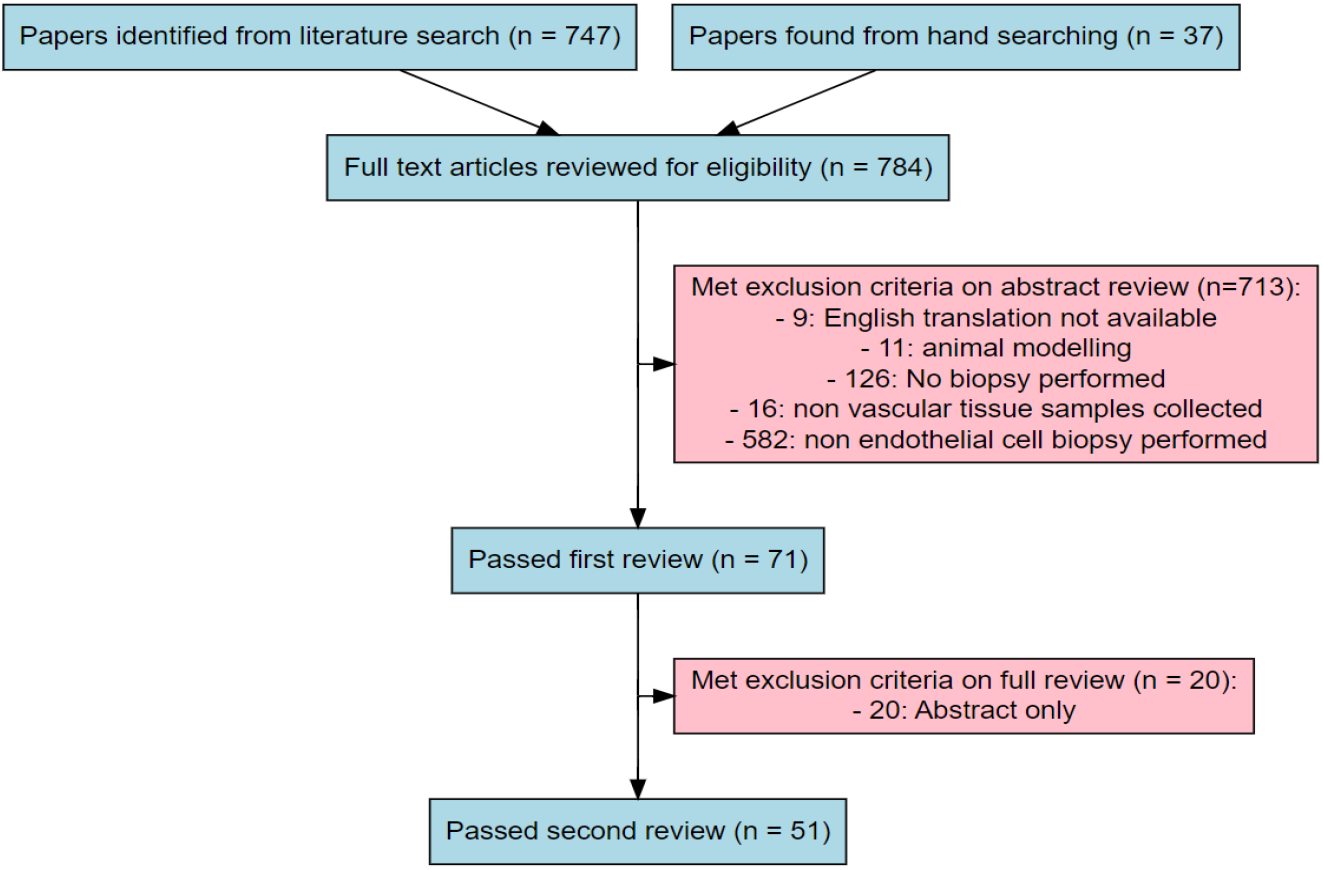
PRISMA diagram visualising the search strategy and inclusion/exclusion criteria.

### Study participant characteristics

Study participant characteristics were available from 50 studies included in this review. 24 studies collected EC from healthy participants only and investigated the effect of sedentary lifestyle, sex, age, smoking and weight on the endothelium (12–33). 26 studies collected EC from patients, 14 of which also collected samples from healthy participants as controls. Endothelial cells (ECs) from patients were studied in five main areas, as summarised in Table I.

**Table I:** Frequency of studies and number of participants according to characteristics of cohort.

|  | **Number of studies** | **Number of participants** |
| --- | --- | --- |
| Healthy participants only | 24 | 1,679 |
| Cardiovascular | 9 | 323 |
| Cerebrovascular | 6 | 57 |
| Diabetes mellitus (T2) | 6 | 202 |
| Obstructive sleep apnoea | 4 | 271 |
| Arteriovenous fistulae | 1 | 10 |

### Biopsy site

Endothelial cells (EC) were successfully isolated from ten different anatomical sites, see Figure 1. The most frequently sampled location was the superficial forearm veins via access at the cubital fossa, as described in thirty studies (12,13,16,17,20,22,25–28,31,32,34–51). Among these, eleven studies reported the number of EC collected from this site, with two studies presenting separate results from forearm vein samples (13,52,36–38,41,50,20,23,24,26). Weighted means analysis indicated that sampling from forearm veins yields an average of 1,207 ± 1,138 EC per patient.

Samples were also obtained from major arm arteries: three studies sampled the brachial artery (21,30,53) and four studies sampled EC from the radial artery (29,33,36,54). Data on EC yield from the brachial artery was not available, and only one study reported results for the radial artery, which yielded 752 ± 107 EC per patient. Additionally, one study obtained samples from the brachiocephalic vein fistulas, (24 ± 2 EC per sample) (46). EC were also collected from central arteries: the aortic arch (55) and another from coronary arteries (56), yielding 96 EC per sample. One study included the internal carotid artery which yielded 248 ± 37 per sample(11). In terms of major intracerebral vessels, one study sampled EC from the middle cerebral artery (57) and two studies sampled cerebral aneurysms (58,59) with both providing yield data and success rates. A weighted mean showed an average of 17 ± 10 EC per sample. Finally, EC were sampled from major peripheral arteries. Five studies investigated the iliac artery (11,58–61), with three providing information on yield. A weighted mean analysis showed an average of 101 ± 104 EC per patient. Finally, one study sampled the femoral artery which yielded 23 EC per patient (59). This is further outlined in Figure 2.

**Figure 2:**
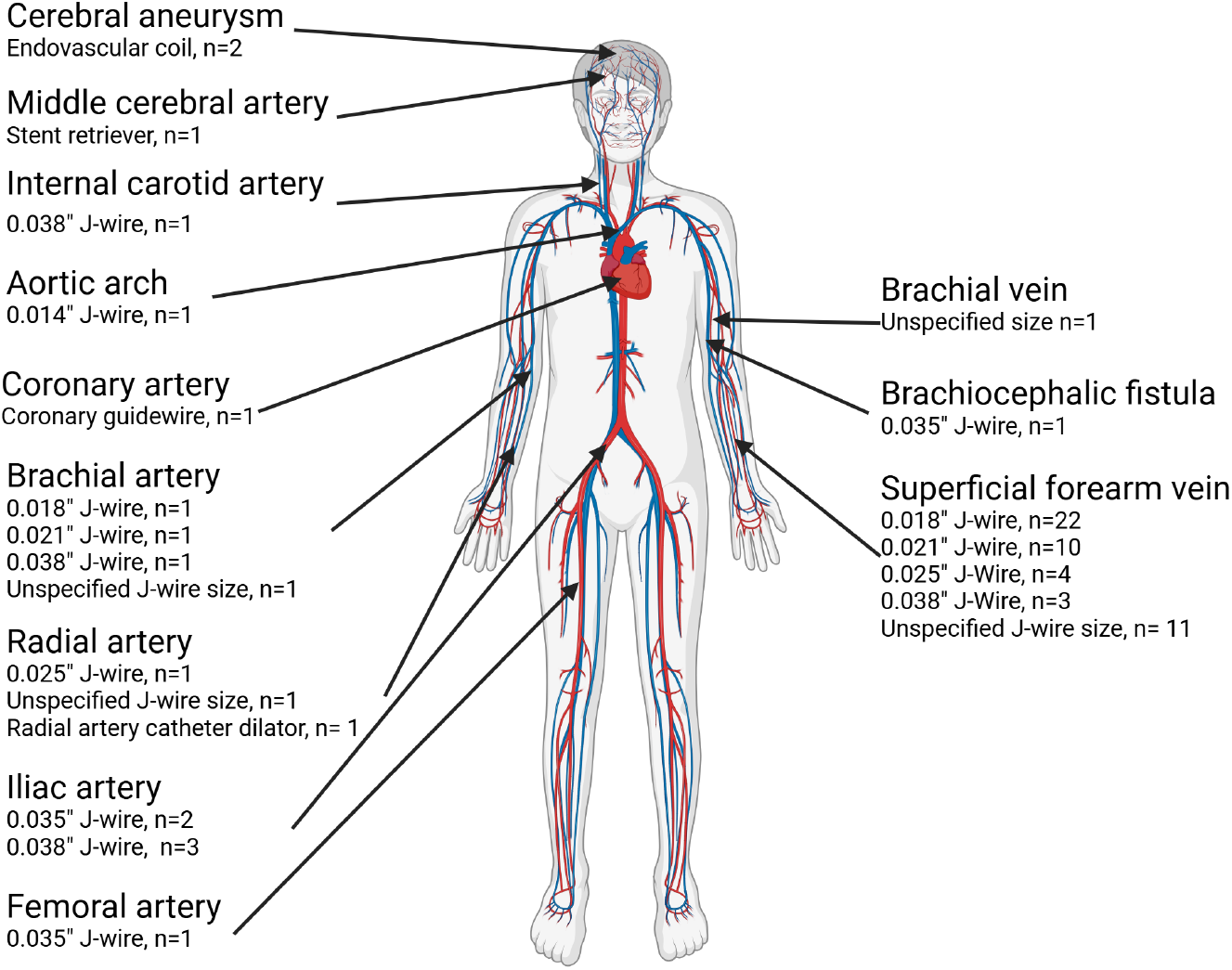
Distribution of sampling sites and guidewire used for endothelial cell biopsy.

### EC biopsy device

Four different interventional devices were used to successfully obtain EC. The most frequently used device was a “J-shaped” guide wire described in 46 studies summarised in Table II. Other devices included endovascular coils (58,59), stent retrievers (57) and radial catheter sheaths (54).

**Table II:** Size of “J-shaped” guidewire used with weighted mean analysis or yield.

| Guidewire size | Number of studies | Number of studies that reported yield | Weighted mean yield | References |
| --- | --- | --- | --- | --- |
| 0.014" | 1 | 1 | 96 | (56) |
| 0.018" | 23 | 1 | 50 | (14–19,21,22,24,26,28,34–36,39,42–45,48,49,53) |
| 0.021 | 8 | 6 | 471 ± 144 | (15,16,18,21,26,36,37,53) |
| 0.025" | 3 | 1 | 4,214 | (23,29,31) |
| 0.035" | 4 | 5 | 51 ± 57 | (46,58–60) |
| 0.038" | 6 | 3 | 181 ± 128 | (11,12,15,18,21,53,61) |
| Multiple sizes | 8 |  |  | (15,16,18,21,26,36,53) |
| Unspecified | 15 | 6 | 1558 ± 503 | (12,13,20,25,27,30,33,38,40,41,47,50–52,55) |

The “J-shaped” guide wire method involves passing a metal J-wire into a blood vessel through a range of intravenous catheters and sheaths. Using weighted means analysis, the yield of EC varied between 50 and 4,214 ECs per patient as summarised in Table II. Linear regression analysis did not reveal any relationship between size of guidewire and yield.

0.014”(n=1) yielded a mean of 96 EC per patient, 0.018” (n=1) yielded a mean of 50 EC per patient, 0.021” (n=6) guidewires yielded a mean of 471 ± 144 EC per patient, 0.025” guidewires (n=1) 4,214 EC per patient, 0.035” guidewires (n=5) yielded a mean of 51 ± 57 per patient, 0.038” guidewires (n=3) yielded a mean of 181 ± 128 per patient. Studies that did not define the size of guidewire (n=6) yielded a mean of 1,558 ± 503 EC per patient.

Endovascular coils applied during the treatment of aneurysms were used twice to obtain EC. This technique involved using telescopic guides and microcatheters matched to patient anatomy to retrieve EC. If deemed safe by the operating clinician, the coil was removed for EC sampling with a second coil implanted for treatment. Both studies provided information on the number of EC obtained. Weighted mean analysis give a mean of 17 ± 10 EC per coil. Stent retrievers used during standard practice and radial catheter dilator sheaths used during standard practice were reported.

### Identification and characterisation of EC

Several biomarkers were used for the identification and characterisation of EC. The most frequently used biomarker was von Willebrand factor (vWF), which appeared in 26 studies. 24 of these used vWF as the sole biomarker, while two studies combined it with other EC markers. In 25 of these studies, immunofluorescence of measurement of vWF was used to identify EC. Additionally one study used laser capture microdissection (LCM) to identify EC (12,15,16,18– 21,24,26,27,30,34–37,41,42,44,44,45,48,50,52–55,60). Other frequently used markers include CD144 which was in 6 studies as the sole biomarker and 9 studies in total. Immunofluorescent measurement of CD144 was used four times to characterise EC with FACS used twice and MACS used once (13,25,29,31–33,40,47,51). CD146 which was used in four studies as the sole biomarker and ten times in total. MACS enrichment of CD146 was used six times and FACS used four times to (14,23,38,40,43,46,55,56,58,60), CD31 which was used once as the sole biomarker and eight times in total(31,46,51,57–61). FACS identification of CD31 was used five times to identify EC and LCM used once. CD34 which was used in five studies and always in combination with other markers (23,46,58,59,61) and only used to characterise EC using FACS. CD105 which was used in five studies (23,46,58,60,61), also always in combination with other markers and only used in FACS identification of EC. Less commonly used markers include endothelial nitric oxide synthase (eNOS), which was employed in only three studies (28,39,54), all of which used immunofluorescence to identify EC. See figure 3 for details on combinations of biomarkers.

**Figure 3:**
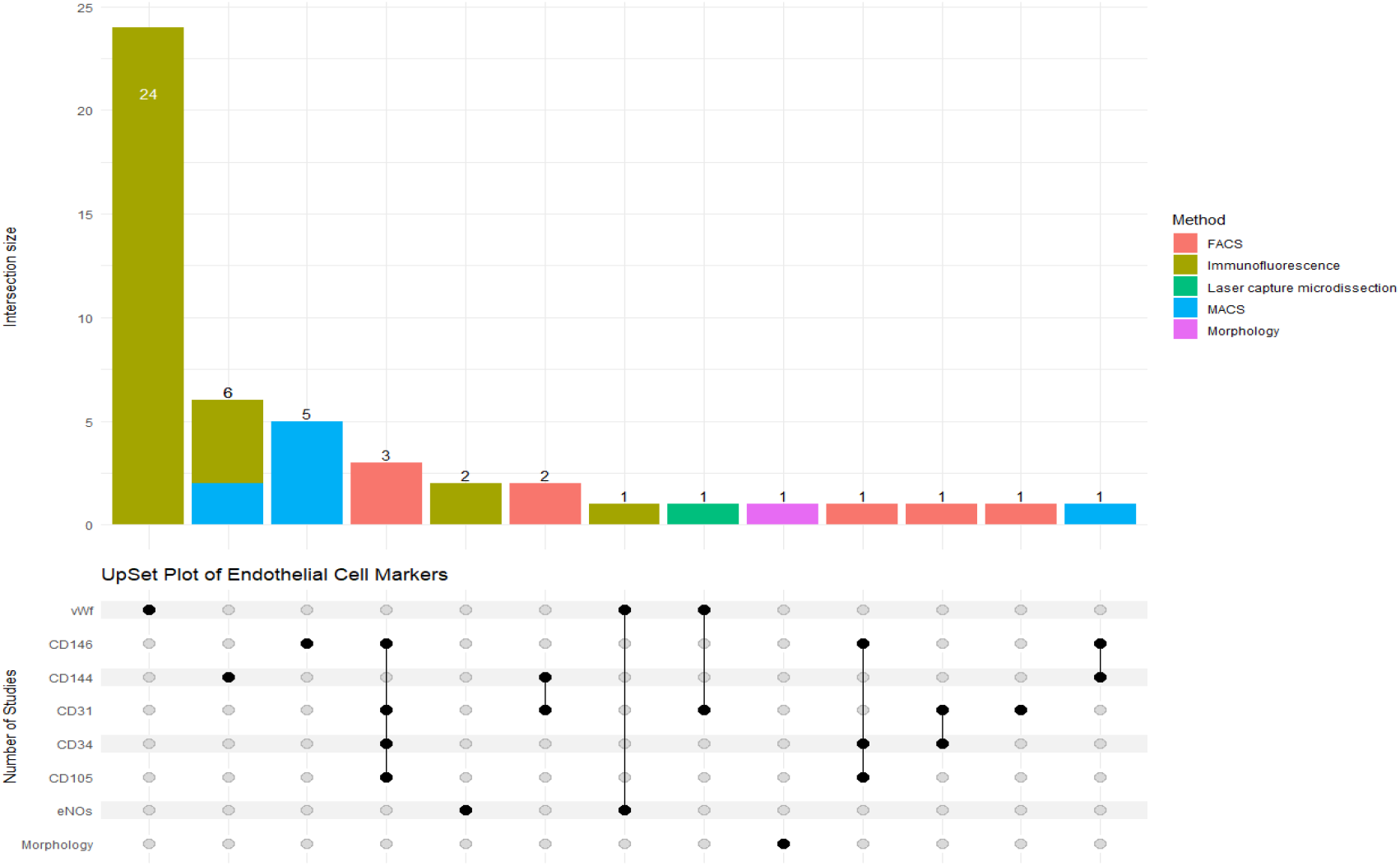
Frequency and combination of biomarkers and identification techniques used for characterisation of EC.

### EC biopsy yield

16 studies provided information on the yield of endothelial cells obtained from biopsy. Of these four studies provided full information on the average number of EC obtained including standard deviation (11,37,46,58,60) and 12 studies only provided only an average. A meta-analysis was not feasible due to the variability in reporting of EC yield and the heterogeneity of methods used. However, when the data from these studies were combined using a weighted mean approach, the average yield of endothelial cells obtained was 1,058 ± 893 EC per biopsy.

### Depth of wire insertion

The depth of wire insertion was reported in 21 studies, all of which exclusively used J-wires. The most common insertion depth was 10 cm, reported in 11 studies (12– 14,19,20,24,38,40,41,43,50). One study inserted J-wires to a depth of 7.5 cm in two different scenarios (60). Three studies reported an insertion depth of 5 cm (31,51,61). Five studies inserted to a depth of 4 cm (15,21,26,29,53). Lastly, one study inserted wires to a depth of 1 cm (46).

### Number of wires used per biopsy

The number of wires used per biopsy ranged from 1 to 5. In 31 studies, only one wire was used (11,16–18,22,25–30,32,34–37,39,42,44–46,48,49,52,54–59,61). Six studies utilised two wires (15,19,21,24,53,60) but only one study reported yield data (60) in two different scenarios. In eight studies, three wires were used (12,13,20,38,40,41,47,50). Two studies used five wires EC wires (14,43).

### Side effects or complications

Only 6 of the 43 studies specifically refer to adverse event data related to the biopsy procedure. Of these, 4 mentioned no adverse events and only 2 reported adverse events. The first study documented that all participants (n=24) felt minimal pain and discomfort during the procedure, with only 1 patient developing superficial phlebitis which was treated conservatively and resolved. The second study documented that only 1 patient out of 24 developed phlebitis. It was described as painful but benign and required treatment with 2 days of non-steroidal anti-inflammatory medications.

### Enrichment

The sample obtained from the wire required processing to maximise the proportion of EC through an enrichment process. Non-endothelial cells that need to be excluded include erythrocytes and leucocytes. EC were enriched from samples using either Fluorescence-Activated Cell Sorting (FACS), magnetic bead enrichment or laser capture microdissection. FACS was used in n=9 studies (23,24,46,47,57–61). Magnetic beads were used in n=9 studies, with CD144 employed in n=3 studies and CD146 in n=6 studies (14,32,38,40,43,47,52,55,56). Laser capture microdissection was used in n=1 study (60).

### Success rate

The success rate was defined as the number of samples collected with an EC yield high enough for planned downstream analysis as a proportion of the number of patients who underwent the biopsy technique. This was reported in 32 studies (11,13,14,22,25– 39,41,43,45,46,49,51,52,54,55,57–60), and 19 did not document success rates (12,15–21,23,24,40,42,44,47,48,50,53,56,61). Success rates ranged from 41% to 100%. Weighted analysis indicated an overall average success rate of91 ± 15%.

### Downstream analysis

A range of techniques were used to analyse EC isolated from patients. Most commonly immunofluorescence which was used in 36 separate studies (12,13,15–22,24–30,32–37,39– 42,44,45,47–50,52–54,56). The lowest reported number of EC required for successful analysis was 20 EC (22,34,35,39,45).

Several transcriptomic techniques were reported including, qRT-PCR used in 8 studies (13,23,40,40,43,46,55,60) and sc-PCR used in 3 studies (11,58,61). The lowest reported number of EC required for successful analysis was 23 EC (46). RNA-seq used in 2 studies (52,59) and the lowest reported number of EC required for successful analysis was 1,000 EC (52). Other techniques used in microarray RNA analysis 1 study (14), FACS and flow cytometry in 5 studies (23,31,51,57,61), Western blotting was used in 3 studies (34,36,42) and phage library construction used in one study (38).

## Discussion

This systematic review provides a comprehensive synthesis of the literature on ECBx, summarising findings from 51 human studies. Our results confirm that ECBx has been successfully performed in diverse patient populations, including healthy individuals and those with cardiovascular, cerebrovascular, and metabolic diseases. The findings highlight considerable variability in ECBx protocols, particularly in device selection, insertion sites, and procedural techniques. The J-wire (0.018”) inserted into a forearm vein emerged as the most frequently used method; however, variations in guidewire size, insertion depth, and vascular access sites were evident. While this flexibility allows adaptation to different patient populations, it also underscores the lack of standardised protocols, which may limit comparability across studies and impact reproducibility. Establishing consensus guidelines for optimal wire selection, insertion depth, and sampling technique could improve procedural consistency and clinical applicability. Only 16 studies quantified EC yield, giving a weighted mean of 1,058 ± 893 ECs per biopsy, with a 91% procedural success rate. While these results suggest that ECBx is a reliable method for obtaining ECs, the large yield variability may reflect the technique’s inconsistency.

EC characterisation primarily relied on traditional biomarkers, vWF, CD31, CD144, and CD146 being the most frequently used markers. Although these biomarkers are well-established, their frequent co-expression means that reliance on them should be carefully considered(62). The emerging use of next-generation techniques, such as scRNA-seq, presents an exciting opportunity to refine endothelial profiling and uncover novel pathways. Incorporating these high-resolution approaches could enhance our understanding of vascular pathophysiology and facilitate the development of personalised vascular therapies.

Some studies reported using up to five guidewires to increase EC yield, raising concerns about potential endothelial trauma and vascular perturbation. While safety reporting was inconsistent, six studies that documented complications noted no serious adverse events, though minor complications were relatively common. This suggests ECBx is a low-risk procedure, but the absence of systematic safety reporting is concerning. Future research should assess whether multiple passes introduce iatrogenic endothelial activation, potentially confounding analyses, and establish standardised safety monitoring. Given the increasing interest in precision medicine, ensuring a comprehensive safety profile for ECBx is critical before broader clinical adoption.

To gain greater understanding of endothelial cell biology and vascular medicine in health and disease in humans, it is useful to acquire endothelial cells for detailed characterisation and *ex vivo* experimentation. Many researchers rely on animal models, the acquisition of circulating EC from humans, or isolation of EC from human tissues like the umbilical cord (63,64). However, these approaches face important limitations: animal models often lack translatability, circulating ECs are not organ-specific and are not accepted as traditional EC, and surgical tissue is difficult to obtain (8,65). An alternative approach is to use commercially available human ECs derived from specific vascular beds, such as human umbilical vein, human coronary artery, or human dermal microcirculation. While these models are easily established and manipulated, they come with significant limitations, primarily that they are not patient or disease specific. Other limitations include susceptibility to culture conditions, variability due to cell passage number, and the loss of paracrine signalling from neighbouring cells. Furthermore, traditional models fail to replicate the flow and shear stress conditions that ECs experience in vivo and may not fully represent the heterogeneity of vascular beds across the body (66–68). To address these limitations, 3D spheroid cultures and bioreactors have been developed, offering a more physiologically relevant environment for studying EC behaviour (69–73). However, these models have limitations, including cost and the need for highly specialised training to operate, and they often use animal cells due to the challenges of obtaining human tissue. ECBX offers a promising alternative approach to the direct isolation of ECs from patients (potentially serially) affected by specific conditions using endovascular endothelial cell biopsies techniques.

The strengths of this review include the comprehensive overview of the published literature, highlighting the various patient cohorts and study settings used. Data was collected from multiple research groups over the course of 25 years since the initial publication. The data also illustrates the evolution of the ECBx technique and its potential applications, matching the progression of molecular analytics.

The main limitations of the included studies were the variation in patient characteristics (healthy volunteers or patients with specific pathology), technical aspects (choice of wire, number of wires, depth of wire insertion, location of wire insertion), and analytical techniques applied (dissociation buffer, enrichment technique, choice of final analysis). The reporting of these variables was inconsistent and showed significant variability, likely due to ECBx and reporting of yield not being the primary endpoints of the study. Therefore, statistical analysis to obtain insights to define optimal technique to assist researchers was limited. For example, it was possible to compare EC yield per wire between different depths of insertion, but the interpretation of these findings would be substantially limited by other differences between the studies. Therefore, the main analyses included were limited to weighted means of individual variables, without more complex analyses being reported.

In addition to deliberate instrumentation of a vessel, novel approaches that “piggy-back” onto clinically indicted vascular procedures such as stent retrievers, radial catheter sheaths, and endovascular coils (which are typically applied in aneurysm treatment) also demonstrated promising outcomes. This highlights the opportunity to use other standard of care methods that use a Seldinger-style (catheter over guidewire) techniques to obtain tissue specific EC. These methods include use of central venous catheters, peripherally inserted central catheters (PICC), or peripheral arterial catheters which are common procedures in modern hospitals(74–76) particularly in acute, critical care and perioperative areas.

All reported studies in patients have been conducted in outpatient settings with patients who have the condition of interest (for example diabetes) but were not acutely unwell. The pathophysiology of the EC response to acute illness in humans has not been described. Acute illnesses like sepsis, trauma, surgery, haemorrhage and anaphylaxis are all associated with hypotension and shock states and EC function is likely to highly relevant to the clinical presentation (77,78).

Researchers reporting cohorts of patient who have had ECBx should include a core dataset that include all pertinent data to permit other researchers to replicate their technique. This should include a comprehensive description of the guidewire(s), and how it was used, as well as providing details on EC yield and how the EC were enriched and subsequently analysed. The protocol and standard operating procedure for ECBx should be comprehensive and included in any publications as supplementary materials. In accordance with Good Clinical Practice (International Council for Harmonisation of Technical Requirements for Pharmaceuticals for Human Use), adverse event data should be collected, and it is highly desirable that it is also reported.

In conclusion, ECBx can be safely obtained from volunteers or patients with a low risk of complications and with sufficient yield to allow a range of advanced analytical techniques, including multiplex flow cytometry or RNA-Seq. Use of ECBx offers the potential to gain new mechanistic insights into the pathophysiology of diseases that affect the endothelium and could ultimately contribute towards the development of new and personalised therapies.

## Data Availability

All data produced in the present work are contained in the manuscript

## Ethical Approvals

No human studies were carried out by the authors of this review. No ethical approvals were required for this review. The review was registered on PROSPERO (CRD42022278551) on 21^st^ February 2022.

